# SARS-CoV-2 antibody seroprevalence in cancer patients on systemic antineoplastic treatment in the first wave of the COVID-19 pandemic in Portugal

**DOI:** 10.1101/2022.01.31.22270178

**Authors:** Gonçalo Fernandes, Paulo Paixão, Laura Brum, Teresa Padrão, Jorge Correia, Joana Albuquerque, Catarina Pulido, Mónica Nave, Teresa Timóteo, Tânia Rodrigues, Filipe Costa, José Luís Passos Coelho

**Affiliations:** Serviço de Oncologia Médica, Hospital da Luz Lisboa; Serviço de Patologia Clínica, Hospital da Luz Lisboa; SYNLAB Portugal

**Keywords:** COVID-19, SARS-CoV-2, serology, seroprevalence, oncology, cancer

## Abstract

At the time of the first wave of the COVID-19 pandemic cancer patients were considered to be at high risk of serious illness and had a higher exposure risk since they needed frequent and non-deferrable hospital visits. Serologic tests were not routinely used and seroprevalence in this population was unknown.

A single-center cross-sectional study was developed to determine the seroprevalence of anti-SARS-CoV-2 antibodies (Abs) in cancer patients undergoing systemic antineoplastic treatment. One hundred patients were consecutively recruited in a two-week period (6th to 20th May, 2020) and serum samples were tested for the presence of IgM and IgG Abs directed against both spike and nucleocapsid SARS-CoV-2 proteins in two distinct timepoints (at recruitment and four to eight weeks later). IgG positive results were subject to confirmation, in the same serum sample, using two distinct assays.

At the time of the first study visit, no patient had a previously confirmed diagnosis of COVID-19, one reported previous contact with a COVID-19 patient and all had a baseline SARS-CoV-2 negative RT-PCR. Two patients tested positive for SARS-CoV-2 IgG in the first study visit, which was not confirmed in either of the two confirmatory assays. Seventy-two patients were tested at the second study visit, all with negative IgG test. IgM was persistently positive at both study visits in one patient and was positive in another patient at the second study visit, both with negative RT-PCR and serum IgG. No patient tested RT-PCR positive within the study timeframe.

No evidence of prior or acute SARS-CoV-2 infection was documented in this cohort of cancer patients undergoing systemic treatment and no additional exposure risk was documented compared to general population seroprevalence studies. The study was inconclusive regarding the role of SARS-CoV-2 serology in cancer patients in the early phase of the pandemic. This study did show that, with adherence to recommended preventive measures, it was safe to maintain systemic cancer therapy.

## Introduction

The majority of patients with COVID-19 develop antibodies (Abs) against SARS-CoV-2.[1] While the gold standard for acute COVID-19 diagnosis remains the detection of SARS-CoV-2 virus in respiratory tract swab specimens by RT-PCR[2], serological tests detecting Abs, IgG and IgM, may identify patients that have been infected in the past, including prior asymptomatic infections, and can be used to measure herd immunity to the disease.[3]

Available scientific evidence at the time of the first COVID-19 pandemic wave indicated a worse prognosis of the disease in cancer patients, with early reports from China showing that the overall case-fatality rate was 2% in general population and 5,6% in patients with preexisting cancer[4], and in one cohort the 30-day mortality rate reached 29%.[5] Since April 2020, Portuguese National Health Authority recommendations determined that molecular nucleic acid amplification tests for SARS-CoV-2 detection in upper respiratory tract swab specimens should be performed prior to each treatment cycle in cancer patients undergoing chemotherapy, even in asymptomatic patients.[6] Serologic tests were not routinely used at that time and there was scarce available data on seroprevalence in this patient population.

Cancer patients needed non-deferrable hospital visits, both for evaluation and for treatment, despite the general population “stay at home” practice. We hypothesized that these patients could be at a greater exposure risk, and we developed a cross-sectional study to determine the seroprevalence of anti-SARS-CoV-2 antibodies (IgM e IgG), at two distinct timepoints during the first wave of COVID-19 pandemic, in cancer patients (solid tumors or hematological cancer) undergoing systemic antineoplastic treatment in our Oncology Unit.

## Methods

### Study design

The study included two outpatient visits. A two-visit design was used to minimize the risk of false-negative results associated with testing during early disease and the subsequent possibility of and undetectable level of specific antibodies (window period).[7]

On day one (first study visit), eligible patients were recruited to the study and written informed consent was obtained. An extra blood sample was collected for serological assays (anti-SARS-CoV-2 IgM and IgG) at the same moment of blood collection for routine scheduled tests (no additional venous puncture was needed) and patients were asked to fill in two paper questionnaires (symptoms and epidemiology).

On study day 29 to 57 (4- to 8-week period after first visit) patients that remained on active treatment with chemotherapy or immunotherapy at our Unit had a follow-up anti-SARS-CoV-2 serological test (second study visit). Blood collection was performed at the same moment of blood collection for routine tests and no additional puncture was needed. Again, patients were asked to fill in two questionnaires (symptoms and epidemiology).

Fourteen days after each study visit patients received a follow-up phone call for assessment of symptoms attributable to COVID-19 and / or diagnosis of COVID-19 in the previous 14 days.

The results of the serological tests were not known at time of treatment administration. There was no sufficient evidence on interpretation of these results and therefore they didn’t influence patient management.

### Study sample

This study was designed to include 100 patients, a convenience sample that reflects the expected accrual of patients in a two- to three-week period, considering the number of patients treated at our Medical Oncology Unit fulfilling the study inclusion criteria. Recruitment started on May 6^th^, 2020 and was planned to continue until patient 100 was included.

Inclusion criteria were: 18 years-old or older; ability to understand and sign a written informed consent; diagnosis, documented in the electronic medical record (EMR), of one or more solid or hematologic malignancies, regardless of the clinical or pathological staging; treatment with chemotherapy or immunotherapy (or its combination, with or without other concomitant therapies, including radiation therapy), administered on an outpatient basis, orally or intravenously; SARS-CoV-2 RNA molecular detection test in a respiratory sample (oropharynx and deep nasopharynx) within maximum 30 days prior to study enrolment. Patients were excluded in case of a previous diagnosis of symptomatic COVID-19, presumed or confirmed, if signs or symptoms possibly related to SARS-CoV-2 infection were present in the previous 14 days or if they were participating in a clinical trial.

### Laboratory procedures

IgM and IgG Abs directed against both spike (S) and nucleocapsid (N) SARS-CoV-2 proteins were analyzed by MAGLUMI 2019-nCoV IgG and IgM (Snibe, Shenzhen, China) chemiluminescense assay (specificity 97%). In addition, IgG positive results were subject to confirmation, in the same serum sample, by chemiluminescence assays IgG Abbott Architect SARS-CoV-2 (Abbott Diagnostics, Chicago, USA; specificity >99%) and DiaSorin LIAISON SARS-CoV-2 S1 / S2 (DiaSorin, Saluggia, Italy; specificity 98,5%), respectively detecting antibodies specific for SARS-CoV-2 N and S proteins.

In chemiluminescence-based assays the detection of IgG or IgM is based on double-antibodies sandwich immunoassay. Recombinant antigens rN and rS were conjugated with fluorescein isothiocyanate (FITC) and immobilized on the anti-FITC antibody conjugated magnetic particles. Alkaline phosphatase conjugated human IgG/IgM antibody was used as the detection antibody. An automated magnetic chemiluminescence analyzer was used to read the measured values of chemiluminescence. Results of these tests are presented as arbitrary units.[8] Manufacturer defined cutoffs for positivity were greater than or equal to 1.00 arbitrary units per mL (AU/mL) for the MAGLUMI 2019-nCoV IgG and IgM, 1.40 signal/cut off index (Index (S/C)) for the IgG Abbott Architect SARS-CoV-2 and greater than or equal to 15.0 AU/mL for the DiaSorin LIAISON SARS-CoV-2 S1 / S2.

SARS-CoV-2 RNA molecular detection tests in respiratory samples (henceforth designated by RT-PCR) were routinely performed according to standard clinical practice and not part of study procedures. Therefore, different laboratory methods and timings may have been applied.

### Statistical considerations

In this study, seropositivity of a sample was determined according to the cutoff value defined by the manufacturer of the serological test, with seroprevalence defined as the proportion of seropositive individuals in the sample.

Continuous variables are described as mean ± standard deviation or median and interquartile range (IQR), depending on the distribution underlying the data. Normality underlying the data was assessed using the Shapiro-Wilk test. For categorical variables the results are presented as n (%).

## Results

One hundred patients were consecutively enrolled during a period of two weeks (6th to 20th May 2020) at the Oncology Department of Hospital da Luz Lisboa (Figure 1).

**Figure 1.**
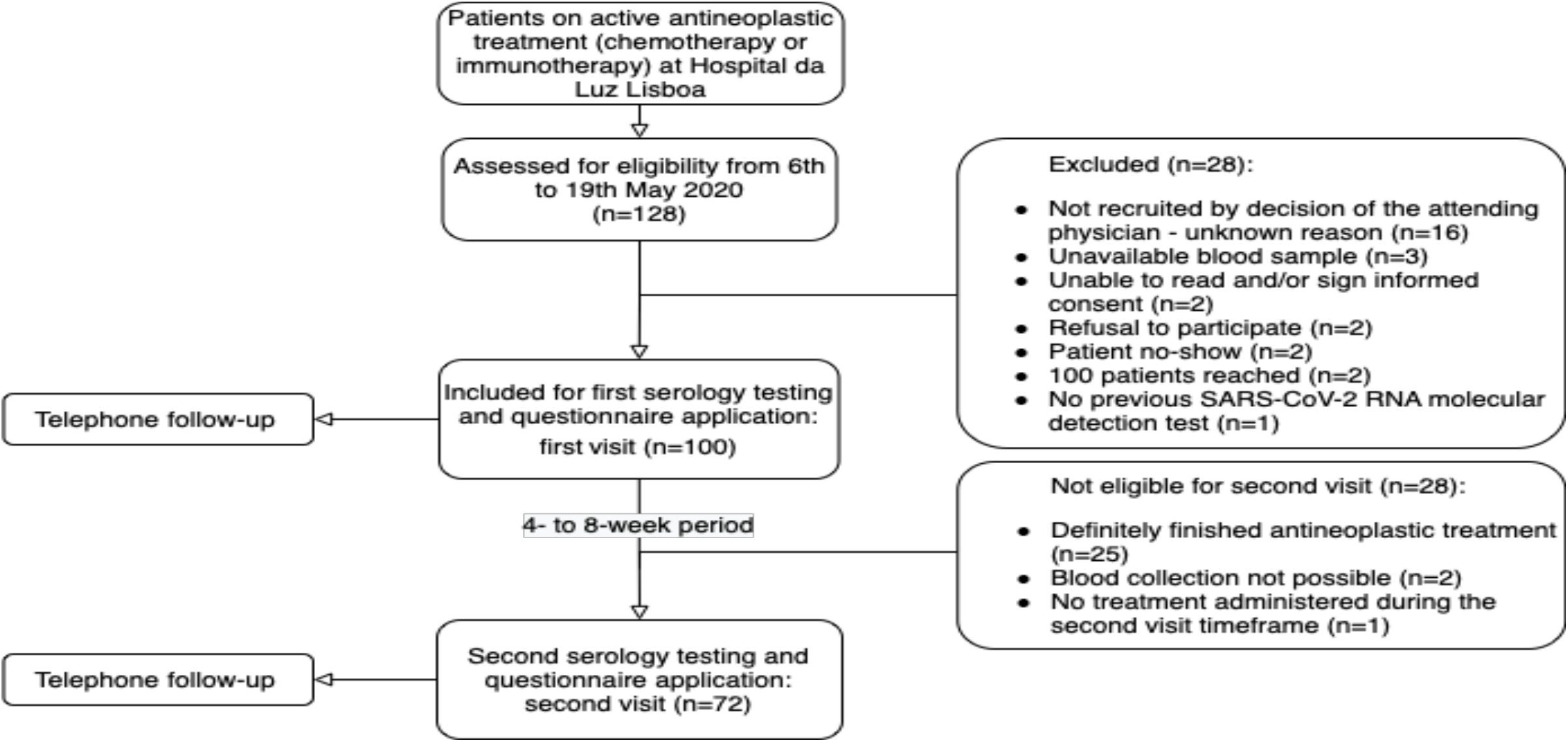
Study design and patient selection flowchart.

Median age was 64 (interquartile range (IQR) 16), the majority were female (61%), and 95% had ECOG Performance Status of zero or one. The majority of malignancies were carcinomas (90%), with a vast range of primary locations, including head and neck, digestive system, lung, breast, genito-urinary, central nervous system and hematological. At time of first visit, 56% of patients were on systemic treatment with curative intent and 44% on palliative treatment (23% first line, 21% second line or subsequent). Ninety seven percent were receiving chemotherapy, alone or combined with anti-HER2, anti-VEGF, anti-EGFR, anti-CD20 or proteasome inhibitor, and 3% were being treated with anti-PD-1/PD-L1 alone. Detailed baseline characteristics are presented in Appendix 1.

At the time of the first study visit, no patient had a previously confirmed diagnosis of COVID-19 and only one patient reported previous contact with a COVID-19 patient (61 days before first study visit). All patients had negative RT-PCR, a median of four days (IQR 4; min 0; max 29) before study inclusion. Seventeen patients (17%) were tested more than seven days before study inclusion due to: re-scheduling of cancer treatment, without repeating RT-PCR by decision of the attending physician (n=4); urgent change of therapy on the day of inclusion without possibility of repeating the test (n=1); patient refusal to repeat the test (n=2); patient under single agent anti-PD-1/PD-L1 immunotherapy (n=2); reason not documented (n=8).

Seventy-two patients attended the second study visit, that occurred a median of 42 days (IQR 7) after the first one. Most patients were on the same antineoplastic treatment regimen of the first visit (n = 61, 85%), while the remainder were on a different phase of the same sequential treatment protocol (n = 5, 7%) or on a further therapeutic line in the palliative setting (n = 6, 8%). None of these patients had a COVID-19 diagnosis within the study timeframe. Between the first and the second study visits patients were screened according to standard practice by RT-PCR. The median number of SARS-CoV-2 RNA molecular detection tests performed per patient was two (IQR 1, min 0, max 5). All tests were negative for the presence of viral RNA. Thirteen patients (18%) were considered non-tested: in three patients (4%) the tests were performed but not documented on EMR; 10 patients (14%) were not tested (patient refusal (n=1); patient under isolated anti-PD-1/PD-L1 immunotherapy (n=3); unknown reason (n=6)). For the patient with valid test results, median time from last RT-PCR to second study visit date was 2 days (IQR 2,5, min 0, max 56). Reasons for more than seven days between most recent mRNA testing and second visit (n=12) were: delay of treatment, without repeating testing by decision of the attending physician (n=1); patient under single agent anti-PD-1/PD-L1 immunotherapy (n=1); RT-PCR tests performed but results not documented on EMR (n=4); unknown reason (n=6).

To the best of our knowledge, none of the remaining 28 patients not eligible for the second study visit had a COVID-19 diagnosis within the study timeframe.

Patients’ self-reported symptoms experienced within the previous 14 days in both study visits are summarized in Table 1. The most frequently reported complaint was tiredness (66% at visit 1 and 76% at visit two), followed by anorexia, myalgia, arthralgia and dysgeusia. All the reported symptoms were considered by the attending physician to be attributable to an alternative diagnosis other than COVID-19.

**Table 1.**
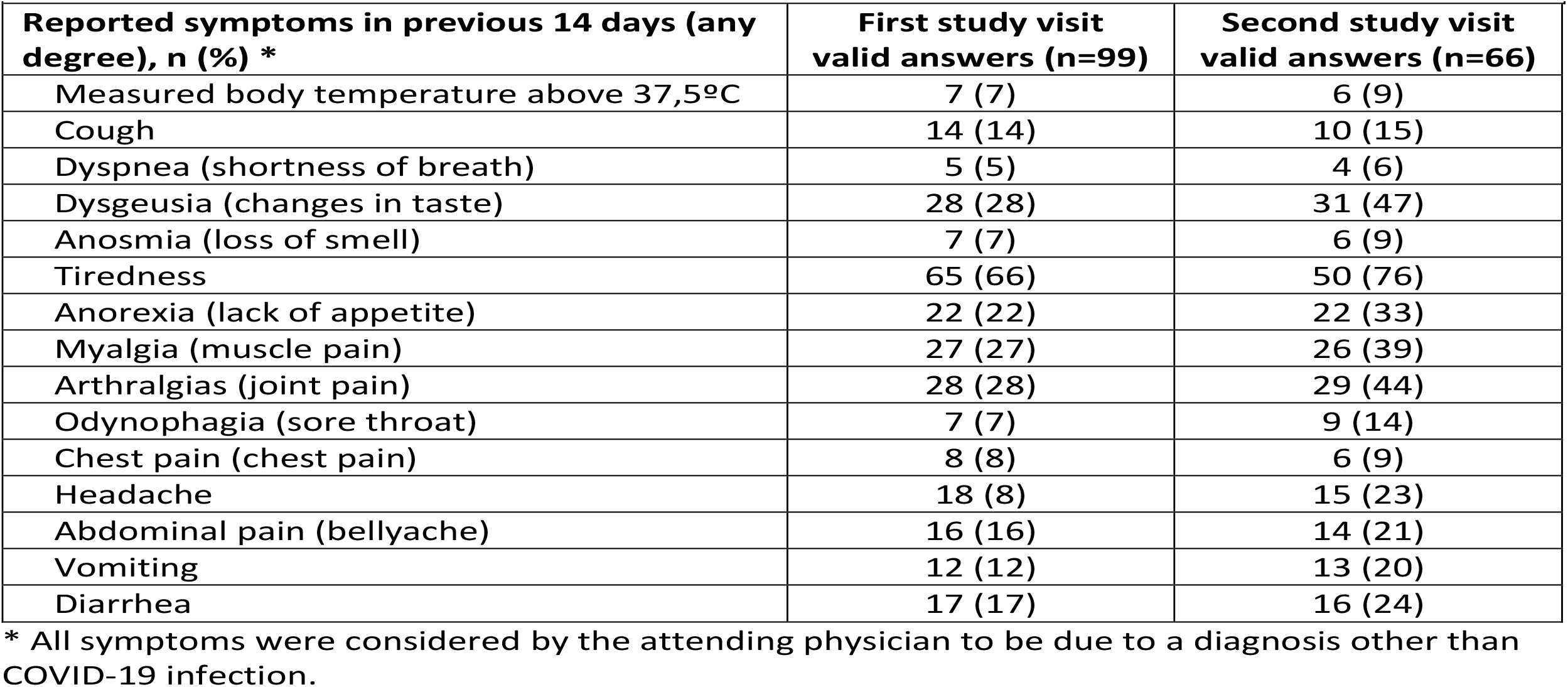
Patient self-reported symptoms.

Results of the serologic assays performed on both study visits are shown in Table 2.

**Table 2.** Serological assay results (MAGLUMI 2019-nCoV (Snibe))

| **Table 2 – Serological assay results (MAGLUMI 2019-nCoV (Snibe))** |  |  |
| --- | --- | --- |
| **Study visit** | **Positive, n (%)** | **Negative, n (%)** |
| **First study visit** |  |  |
| IgM | 1 (1%) | 99 (99%) |
| IgG | 2 (2%) | 98 (98%) |
| **Second study visit** |  |  |
| IgM | 2 (2,8%) | 70 (97,2%) |
| IgG | 0 (0%) | 72 (100%) |

Two patients had a positive SARS-CoV-2 IgG in the first study visit sample, but this result was not confirmed positive in either of the two IgG confirmatory assays (IgG Abbott Architect SARS-CoV- 2 and DiaSorin LIAISON SARS-CoV-2 S1 / S2, respectively detecting N and S proteins) performed in the same serum samples. Both had same day negative SARS-CoV-2 IgM. One of these patients had a same-day negative SARS-CoV-2 RT-PCR test and the other had a similar negative test performed four days prior to the serological test. None of them reported previous contact with a known COVID-19 patient.

The patient that had detectable serum IgM had a same day negative serum IgG and RT-PCR. This patient had no COVID-19 symptoms between first and second visits, maintained IgM positivity and a negative IgG at second visit (56 days after first visit) and was tested RT-PCR negative four times within the same timeframe.

A fourth patient tested serum IgM positive at the second study visit but was IgG-negative and had negative SARS-CoV-2 RT-PCR nasopharyngeal swab the same day.

No patient had a positive serum IgG at second visit. One of the patients with detectable IgG at visit one, tested negative for both IgM and IgG at study visit two. Unfortunately, there was no blood sample collection on visit two from the other IgG-positive patient at study visit one.

Thus, in this population of 100 cancer patients undergoing active cancer treatment we did not document evidence of SARS-CoV2 infection by either mRNA RT-PCR testing or serological testing.

## Discussion

The first cases of COVID-19 were reported from Wuhan, Hubei in China in November 2019. On Jan 31^st^, 2020, Italy reported the first European cases and WHO classified SARS-CoV-2 as a pandemy on March 11th, 2020.[9] As of Dec 8^th^, 2021, more than 266 million cases and 5.27 millions of deaths have been reported.[10] Thus, in April 2020 when this study was planned, Portugal was on lockdown and there were significant doubts on the safety of having patients come to the hospital and regarding the safety of administering cancer chemotherapy to cancer patients with the associated increased risk of infection. As stated before, as of April 2020, the Portuguese Health Authority recommended that cancer patients be tested by RT-PCR prior to the administration of every cycle of active systemic anti-cancer treatment, namely chemotherapy.[6] Thus, it was critical to evaluate the safety of pursuing clinic visits and systemic treatment on cancer patients. Now we know that despite the decision to maintain cancer treatment in most patients, COVID-19 resulted in a delay on cancer diagnosis and an increase in non-COVID-19 related deaths.[11]

Unequivocal evidence of acute or prior infection by SARS-CoV-2 was not documented in this sample of cancer patients undergoing antineoplastic treatment. Thus, to maintain active cancer treatment was shown to be the right clinical decision.

Despite two SARS-CoV-2 IgG-positive patients on the first assay, these results were not confirmed on the two confirmatory assays. Since the beginning of the COVID-19 pandemic, multiple serological assays for anti-SARS-CoV-2 antibody detection have been developed and studied. A meta-analysis and systematic review of studies involving antibody tests to detect SARS-CoV-2 infection summarized evidence from 38 studies involving 7848 individuals and showed very different precision of the available techniques and kits.[8] When our study was designed, the chosen serologic assay was the only one available at our hospital laboratory, with a manufacturers claimed clinical sensitivities for IgM and IgG of 78.65% and 91.21%, respectively, and specificities for IgM and IgG of 97.50% and 97.3%, respectively, and found to be a reliable immunoassay for the measurement of specific IgM and IgG in sera of COVID-19 patients in a validation study.[12]

Given the expected low seroprevalence setting, two different confirmatory assays were used to rule out false-positive IgG results, using the same serum sample. The Abbott Architect SARS-CoV-2 IgG reported a specificity of 99.90% and sensitivity of 100% at day 17 after symptom onset and day 13 after PCR positivity.[13] The DiaSorin LIAISON SARS-CoV-2 S1 / S2 IgG test reported a sensitivity of 97.4% 15 days after disease onset and a specificity of 98.5%; however a study comparing the performance of seven commercially available serological assays, reported a sensitivity of 95.5% and a specificity of 90.5% for this assay.[14] Therefore, we believe that in this 100-patient cohort, the two positive IgG results, without a previous known COVID-19 diagnosis or contact with a COVID-19 patient, not confirmed on the confirmatory serologic assays, were most likely false positive results.

As for the two positive IgM results, no evidence of acute or prior COVID-19 infection was found in these patients and, in accordance with the expected assay performance, false positives cannot be excluded. The patient that remained IgM positive throughout the two study visits, without evidence of IgG seroconversion, suggests an interference with the test results.

Other studies conducted during the first wave of the COVID-19 pandemic showed a low seroprevalence in the general population. A study of 17,368 individuals, in the city of Wuhan, the epicenter of the COVID-19 pandemic in China, showed that the seropositivity varied between 3.2% and 3.8% and progressively decreased in other cities as the distance to the epicenter increased.[15] In Switzerland, a study conducted in Geneva, a region considered to have high prevalence of COVID-19 (5000 reported clinical cases over less than 2.5 months in the population of half a million people) documented a seroprevalence between 4.8% and 10.8%.[16]

In Portugal, a general population study of the prevalence of SARS-CoV-2 specific antibodies performed between May and July 2020 recruited 2301 participants, and reported 30 (1.9%, CI95: 1.1% – 3.1%) IgM-positive and 33 (1.9%, CI95: 1.2% – 3.1%) IgG-positive individuals.[17] Similar to our study, in a tertiary care cancer center in Austria, 84 patients were tested between March 21^st^ and June 4^th^ 2020 with a reported SARS-CoV-2 IgG seroprevalence of 2.4% (95% CI 0.3% to 8.3%).[18]

Overall, the expected general population seroprevalence was low. In Portugal, at the date of the last study sample collection (July 14^th^, 2020), national health authorities had reported 47.051 confirmed COVID-19 cases, approximately 0.5% of the Portuguese population (for a population of ∼10.1 million inhabitants). Among cancer patients, a study reported significantly lower detection rate of SARS-CoV-2 antibodies 15 days or later after development of COVID-19 symptoms and RT-PCR positive testing (versus healthy health care workers), more often undetectable in patients who had received cancer treatments in the month before testing.[19] Since cancer patients are at higher risk of complications from COVID-19[4,5], it is likely that they have higher individual adherence to isolation and protection measures, leading to less exposure to the virus. Protection measures and isolation of cancer patients (e.g., at our hospital the Oncology Unit and Day Hospital were temporarily moved to a separate building to isolate cancer patients from all other patients) also contributed to the safety of maintaining active cancer treatment.

Limitations of this study include its small sample size and the absent second study visit for 28 of the 100 patients. Since RT-PCR was not part of study procedures, rather performed at the discretion of attending physicians, in accordance with standard clinical practice, these tests were performed at a median of four days (IQR 2-6) before serological testing; thus, in theory, acute COVID-19 asymptomatic infections could have been missed on the day of blood collection. At that point in time, low antibody concentrations in the early course of the disease could be below the manufacturer positive cut-off value of the serologic assay.[20] To mitigate this risk, we measured anti-SARS-CoV-2 antibodies at two different time points and, did an extensive review of the patients’ EMR and did follow-up telephone contacts that documented no cases of COVID- 19 infection (symptomatic or asymptomatic) within the study timeframe.

The self-reported symptom questionnaire included symptoms that could be associated with COVID-19 diagnosis but could also be associated with the oncologic disease or its treatment. A Canadian study polled 4240 adults about self-reported COVID-19 experience in March 2020 (early in the pandemic) and found that between 5 and 8% of respondents reported that either themselves or at least one of the household members had possible COVID-19 symptoms.[21] In our study, as expected, cancer patients reported a higher incidence of symptoms that could be COVID-19 related than the previously mentioned study (general population). However, care must be taken when considering self-reported symptoms, as they are subject to limitations and misclassification.

The presented data is not conclusive regarding the role of anti-SARS-CoV-2 serology in the screening of cancer patients for COVID-19 active or prior infection in an early phase of the pandemic. However, given the low incidence of COVID-19 in this study, it raises two questions. First, if the changes or delays in oncologic treatment plan recommended by scientific societies[22], irrespective of local SARS-CoV-2 infection incidence rate, were adequate and what will be the impact on cancer outcomes. Second, the risk of using serologic tests in low incidence settings, given the risk of false positives and the subsequent misinterpretation of the individual immune status.

## Conclusion

COVID-19 was and still is a major public health challenge. However, no additional exposure risk to SARS-CoV-2 was found in this cancer patient cohort compared to results described in general population seroprevalence studies at the time of the first wave of the COVID-19 pandemic. The data reported in this study support the safety of pursuing active cancer treatment despite the restraints of SARS-CoV-2 pandemic. We suggest not delaying cancer diagnosis and treatment and, with the appropriate constraints, we consider that it is possible to deliver cancer treatment safely.

## Data Availability

All data produced in the present study are available upon reasonable request to the authors.

## Ethical considerations

The study protocol was approved by the Hospital da Luz Lisboa Ethics Committee (ref. CES/10A/2020/ME). Written informed consent was obtained from all study participants.

## Acknowledgements

We thank SYNLAB Portugal for the logistic support and collaboration in this work.

### Appendix 1

**Table A.** Baseline characteristics.

| **Table A – Baseline characteristics** |  |
| --- | --- |
| **Baseline characteristic** | **Total sample (n=100)** |
| **Age, years** |  |
| Median | 64 |
| IQR | 16 |
| **Sex, n (%)** |  |
| Female | 61 (61) |
| Male | 39 (39) |
| **ECOG/Zubrod performance status, n (%)** |  |
| 0 | 69 (69) |
| 1 | 26 (26) |
| 2 | 5 (5) |
| **Charlson Comorbidity Index (CCI), n (%)** |  |
| 2 - 3 points | 17 (17) |
| 4 - 5 points | 23 (23) |
| ≥ 6 points | 60 (60) |
| **Smoking status** |  |
| Never-smokers | 44 (44) |
| Former smokers | 39 (39) |
| Current smokers | 6 (6) |
| Unknown | 11 (11) |
| **Malignancy – histology, n (%)** |  |
| Carcinoma | 90 (90) |
| Lymphoma | 7 (7) |
| Central Nervous System | 2 (2) |
| Multiple Myeloma | 1 (1) * |
| **Malignancy – primary site, n (%)** |  |
| Head and neck | 3 (3) |
| Digestive system | 29 (29) |
| Lung | 15 (15) |
| Breast | 35 (35) |
| Genito-urinary | 8 (8) |
| Central Nervous System | 2 (2) |
| Hematological | 8 (8) |
| **Malignancy – Time from diagnosis to first study visit, months** |  |
| Median | 5 |
| IQR | 15 |
| **Systemic treatment intent at time of first visit, n (%)** |  |
| Curative (neoadjuvant/adjuvant or definitive chemoradiotherapy) | 56 (56) |
| Palliative (first line) | 23 (23) |
| Palliative (second line or subsequent) | 21 (21) |
| **Systemic treatment – pharmacological class, n (%)** |  |
| Chemotherapy ** | 97 (97) |
| Anti-PD-1/PD-L1 alone | 3 (3) |
| **Previous surgery (with hospital admission >24 hours), n (%)** |  |
| <60 days | 9 (9) |
| 60-180 days | 25 (25) |
| None or >180 days | 66 (66) |
| **Previous radiotherapy, n (%)** |  |
| <60 days | 5 (5) |
| 60-180 days | 4 (4) |
| None or >180 days | 91 (91) |

\* This patient had a synchronous genito-urinary carcinoma on palliative treatment (not chemotherapy or immunotherapy).
\*\* Alone or combined with anti-HER2 (n=6), anti-VEGF (n=3), anti-EGFR (n=3), anti-CD20 (n=7) or proteasome inhibitor (n=1).

